# Brain and molecular mechanisms underlying the nonlinear association between close friendships, mental health, and cognition in children

**DOI:** 10.1101/2022.11.02.22281840

**Authors:** Chun Shen, Edmund T. Rolls, Shitong Xiang, Christelle Langley, Barbara J. Sahakian, Wei Cheng, Jianfeng Feng

## Abstract

Close friendships are important for mental health and cognition in late childhood. However, whether the more close friends the better, and the underlying neurobiological mechanisms are unknown. Using the Adolescent Brain Cognitive Developmental study, we identified nonlinear associations between the number of close friends, mental health, cognition, and brain structure. Although few close friends were associated with poor mental health, low cognitive functions and small areas of the social brain (e.g., the orbitofrontal cortex, the anterior cingulate cortex, the anterior insula and the temporo-parietal junction), increasing the number of close friends beyond a level (around 5) was no longer associated with better mental health and larger cortical areas, and was even related to lower cognition. In children having no more than 5 close friends, the cortical areas related to the number of close friends revealed correlations with the density of μ-opioid receptors and the expression of OPRM1 and OPRK1, and could partly mediate the association between the number of close friends, ADHD symptoms, and crystalized intelligence. Longitudinal analyses showed that both too few and too many close friends at baseline were associated with more ADHD symptoms and lower crystalized intelligence 2 years later. Additionally, we found friendship network size was nonlinearly associated with well-being and academic performance in an independent social network dataset of middle school students. These findings challenge the traditional idea of “the more, the better”, and provide insights into potential brain and molecular mechanisms.

## Introduction

Late childhood and its transition towards adolescence is a period marked by decreasing parental influence alongside increasing peer influence. It is a period critical for social interaction, during which friendships are especially important^1^. During this period, the social brain is still undergoing significant development, in parallel with changes in social cognition^2^. Evidence suggests that psychiatric disorders often have an onset in adolescence^3^, which may be partly influenced by the concurrent changes in the social environment and brain^4^. Therefore, understanding the relationship between friendship, mental health, and cognition during this period, and the underlying brain mechanisms, is of considerable clinical and public health importance.

It has been well-established that positive social relationships such as close friendships are essential for mental health and cognition in children and adolescents^5–7^. However, it remains unclear whether having more close friends is necessarily better, and if not, whether there is an optimal quantity of close friendships. Cognitive constraints and time resources limit the number of close social ties that an individual can maintain simultaneously^8^. The innermost layer of the friendship group with the highest emotional closeness is around 5 close friends (the so-called Dunbar’s number)^9^. For now, only a few empirical studies have examined the nonlinear association between social relationships, mental health, and cognition. A national investigation reported that adolescents with either too many or too few friends had higher levels of depressive symptoms^10^. Two large-scale studies found that the benefits of social interactions for well-being were nearly negligible once the quantity reached a moderate level^11,12^. A significant U-shaped effect was detected between positive relations with others and cognitive performance^13^. Overall, the assumption of linearity still dominates studies of social relationships, and the effect of the friendship network size at the high end remains largely unexplored.

The social brain hypothesis suggests that complex social selection pressures drive the evolution of brain size: for example, relative neocortex volume correlates with social group size in primates^14^. Both experimental manipulation and free-ranging macaque studies found that social network size could predict mid-superior temporal sulcus volume^15,16^, a region in which neurons respond to socially relevant stimuli such as face expression and head movement to make or break social contact^17^. In humans, a series of studies demonstrated that the medial prefrontal cortex (mPFC, i.e. orbitofrontal [OFC] and anterior cingulate cortex [ACC]), the cortex in the superior temporal sulcus (STS), the temporo-parietal junction (TPJ), amygdala and the anterior insula were involved in social cognitive processes^18,19^. The μ-opioid receptor that is widely distributed in the brain especially in regions implicated in social pain such as the ACC and anterior insula^20^, has been recognized to be crucial to the formation and maintenance of friendships^8^. Variation in the μ-opioid receptor gene (OPRM1) is associated with individual differences in rejection sensitivity^21^. A range of other neurotransmitters including dopamine, serotonin, GABA and noradrenaline may interact with the opioids, and is involved in social affiliation and social behaviour^22^. Dysregulation of the social brain and neurotransmitter systems has also been proposed in the pathophysiology of major psychiatric disorders^23^. While it has been proposed that the association between social relationships and mental health is mediated through changes in the social brain^24^, there is a lack of empirical evidence in late childhood and adolescence.

In this study, based on the existing literature, we hypothesized a nonlinear association between the number of close friends, mental health, and cognitive outcomes. We used data from the Adolescent Brain Cognitive Developmental (ABCD) study^25^, and another independent social network dataset^26^ (N > 23,000 in total; Figure 1a). Two different analytic approaches were used to evaluate the nonlinear relation between friendship quantity (predictor) and mental health and cognition (outcome). A significant quadratic term would indicate the presence of nonlinearity, and then a two-lines test^27^ was conducted to estimate an interrupted regression and to identify the breakpoint (Figure 1b). To explore the underlying neurobiological mechanisms, we further tested the nonlinear association between the number of close friends and brain structure, and then correlated the related brain differences with the density of 8 neurotransmitter systems, and the expression of OPRM1 and κ-opioid receptor (OPRK1) (Figure 1c). Finally, longitudinal and mediation analyses were conducted to uncover the direction and direct association between the number of close friends, mental health, cognition, and brain structure (Figure 1d). We hypothesized that the number of close friends was nonlinearly related to mental health, cognition, and the social brain; and that the nonlinear association between the number of close friends and behavioral measures may be mediated by brain and molecular mechanisms.

**Figure 1.**
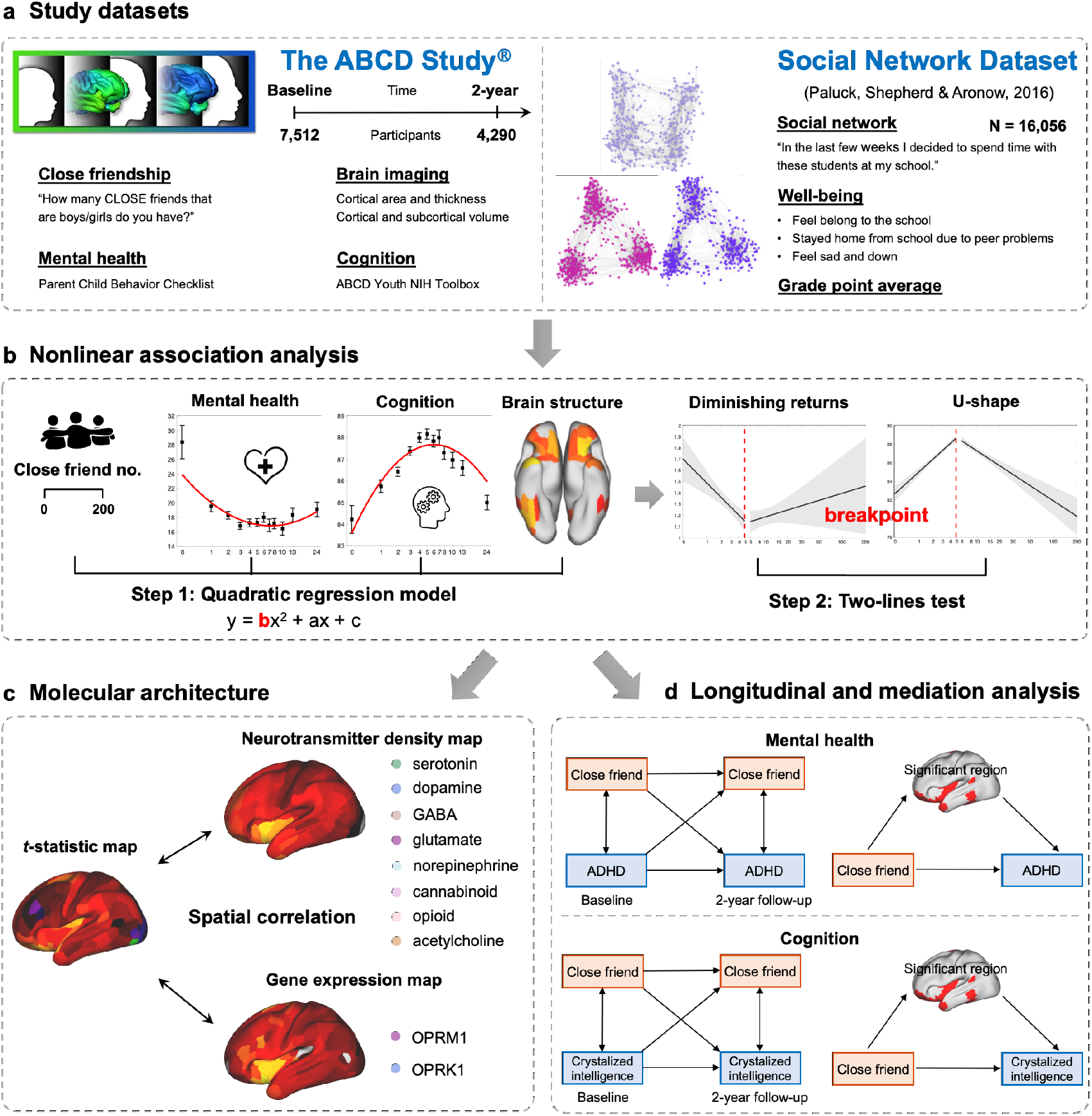
The study workflow. Study datasets and key measures used in the present study (a). A two-step approach to evaluate the nonlinear association (b). The number of close friends is used as the independent variable in quadratic regression models. Once a significant squared term (‘b’) is found, a two-lines test is conducted to estimate the breakpoint. Participants are classified into two groups according to the breakpoint in the following analyses. Correlation of brain differences related to the number of close friends with neurotransmitter density and gene expression level (c). Longitudinal and mediation analysis of the number of close friends, ADHD symptoms, crystalized intelligence and the significant surface areas (d).

## Results

### Demographic characteristics

In the ABCD study, 7,512 participants (3,625 [48.3%] female, aged 9.91 ± 0.62 years) provided self-reported number of close friends, a broad range of mental health and cognitive measures, and quality-controlled MRI data at baseline, and 4,290 of them (2,044 [47.7%] female, aged 11.49 ± 0.66 years) had two-year follow-up data available. Detailed population characteristics are shown in Table 1. In the social network dataset, 16,065 subjects from 48 middle schools (8,065 [50.3%] female, aged 12.00 ± 1.03 years) who had complete key variables were included (Table S1).

**Table 1.** Characteristics of the study population in the ABCD study.

|  | **Baseline (n = 7,512)** | **2-year follow-up (n = 4,290)** |
| --- | --- | --- |
| Age, year | 9.91 (0.62) | 11.49 (0.66) |
| Sex, n (%) |  |  |
| Female | 3,625 (48.3%) | 2,044 (47.7%) |
| Male | 3,887 (51.7%) | 2,246 (52.4%) |
| Race, n (%) |  |  |
| White | 4,169 (55.5%) | 2,612 (60.9%) |
| Black | 931 (12.4%) | 385 (9.0%) |
| Hispanic | 1,500 (20.0%) | 791 (18.4%) |
| Asian | 154 (2.1%) | 89 (2.1%) |
| Other | 758 (10.1%) | 413 (9.6%) |
| Family income | 7.73 (2.35) | 7.83 (2.04) |
| Parental education | 16.87 (2.59) | 17.11 (2.44) |
| BMI | 18.72 (4.11) | 20.35 (4.63) |
| Puberty | 1.75 (0.87) | 2.53 (1.05) |
| Total close friends | 6.26 (9.01) | 6.82 (8.37) |
| Same-sex close friends | 4.78 (6.73) | 4.99 (5.92) |
| Opposite-sex close friends | 1.48 (3.68) | 1.83 (3.66) |

Descriptive statistics are reported as mean ( ± SD) unless noted otherwise.

### Nonlinear association between the number of close friends, mental health and cognition

The number of close friends was significantly associated with 12 out of 20 mental health measures, and 7 out of 10 cognitive scores at baseline (the total F-value of the linear and quadratic terms, *p* < 0.05/30; Figure 2a-2g). For these 19 outcomes except the withdrawn/depressed, all quadratic terms reached significance after Bonferroni corrections (*p* < 0.05/60), and all quadratic models provided a significantly better fit than the corresponding linear models (F = [13.25, 55.53], all *p* < 0.001). For mental health, the greatest effect sizes of the quadratic terms were observed for social problems (*β* = 0.06, *t* = 5.92, *p* = 3.3⨉10^−9^, ΔR^2^ = 0.43%) and attention problems (*β* = 0.08, *t* = 5.83, *p* = 5.8⨉10^−9^, ΔR^2^ = 0.42%). For cognition, the greatest effect sizes of the quadratic terms were observed for total intelligence (*β* = -0.06, *t* = -7.45, *p* = 1.0⨉10^−13^, ΔR^2^ = 0.50%) and crystalized intelligence (*β* = -0.05, *t* = -6.87, *p* = 6.7⨉10^−12^, ΔR^2^ = 0.43%) (Table S2 and Figure S1). The findings were robust with respect to random choice of the siblings (Figure S2).

**Figure 2.**
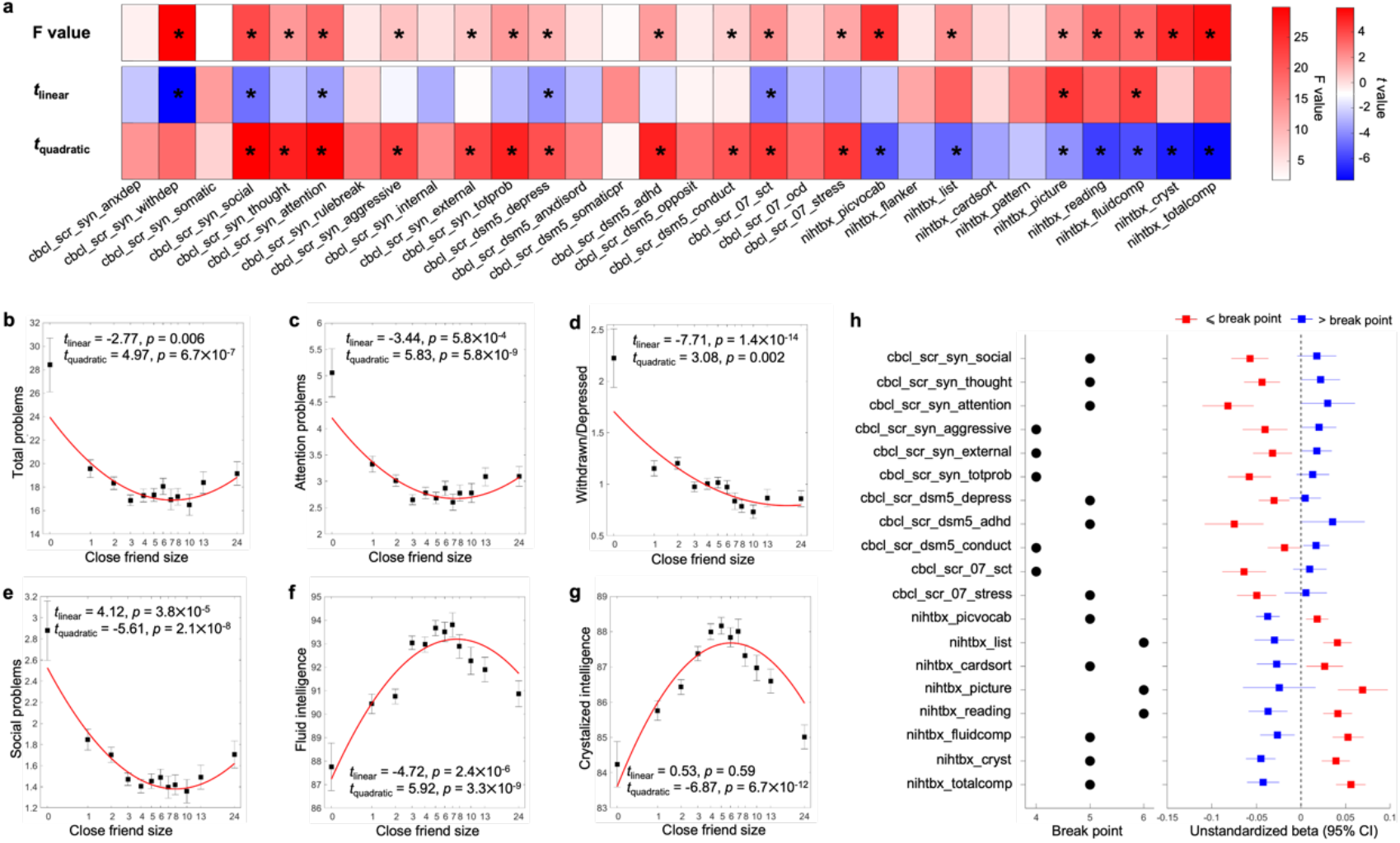
Results of behavior-level nonlinear association analyses in the ABCD study at baseline. Results of quadratic regression models (a). The total F values of quadratic and linear terms, and the *t* values of linear and quadratic terms are reported. An asterisk indicates statistical significance after Bonferroni correction (i.e., *p* < 0.05/30 for F value, and *p* < 0.05/60 for *t* value). Relationship between the number of close friends and the total problems (b), attention problems (c), withdrawn/depressed (d), social problems (e), fluid intelligence (f), and crystalized intelligence (g). The number of close friends is classified into 13 bins. In each bin, the mean (i.e., black dot) and standard error (i.e., error bar) of the dependent variable are shown. The x-axis is in log scale, and the median of the number of close friends in each bin was labeled in the x-axis. The red line is the fitted quadratic model. Results of the two-lines tests (h). The breakpoint and the estimated coefficients with 95% confidence intervals of linear regressions in each group separated by the breakpoint are reported.

The average breakpoint of the number of close friends for the mental health and cognitive outcomes with significant quadratic terms was 4.89 ± 0.66 (Figure 2h). The ideal number of close friends was 5 and the closer to that number, the better for participants’ mental health. Similarly, for cognition the optimal number of close friends was 5 and the further from that number the poorer the participants’ cognition. These nonlinear associations were consistent in males and females (Figure S3). However, the number of same-sex close friends, but not of opposite-sex close friends, was significantly related to mental health and cognition (27 out of 30 measures with a significant F-value after Bonferroni correction), and children with 4.00 ± 0.60 same-sex close friends had the best mental health and cognitive functions (Figure S4). Finally, same analyses were performed using the cross-sectional data collected at 2 years later, and the nonlinear associations of the number of close friends with ADHD symptoms and crystalized intelligence remained significant, with an average breakpoint of 4.83 ± 0.75 close friends (Figure S5).

### The number of close friends was quadratically associated with brain structure

In the ABCD study, the number of close friends was significantly associated with the total cortical area (F = 6.29, *p* = 1.0×10^−3^; Figure 3d), and the total cortical volume (F = 5.80, *p* = 3.1×10^−3^). No significant relationship between the number of close friends and mean cortical thickness (F = 0.62, *p* = 0.54), and total subcortical volume (F = 3.94, *p* = 0.02) was found.

**Figure 3.**
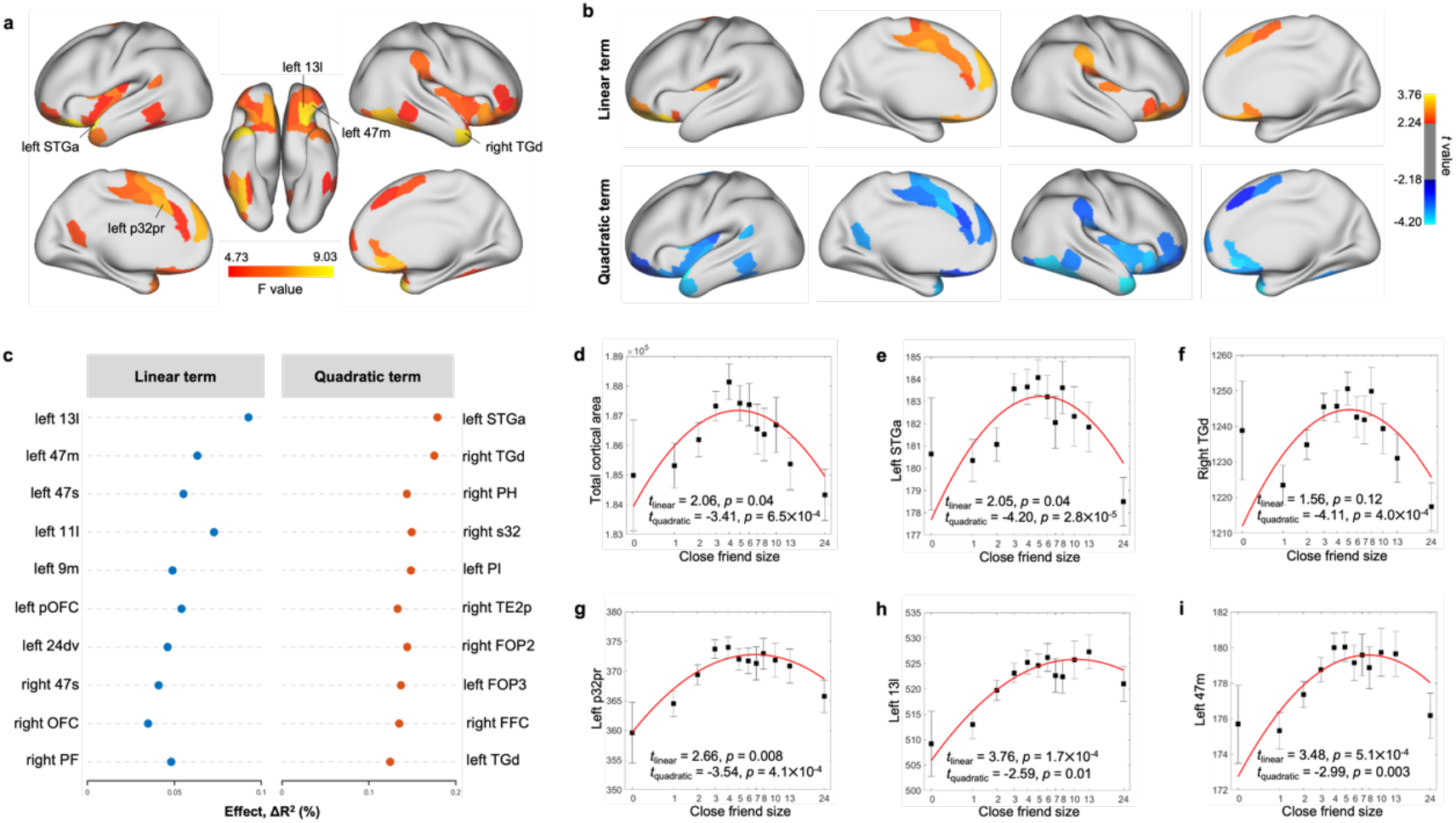
Nonlinear association between the number of close friends and cortical area in the ABCD study at baseline. Cortical areas significantly associated with the number of close friends after FDR correction (i.e., 360 regions) based on the total F values of linear and quadratic terms (a). Cortical areas with a significant linear or quadratic term (b). FDR correction was performed within the significant regions obtained in (a). Top ten regions with the strongest effect sizes of linear and quadratic terms, respectively (c). Relationship between the number of close friends and the total cortical area (d), left STGa (e), right TGd (f), left p32pr (g), left 13l (h), and left 47m (i). The number of close friends is classified into 13 bins. In each bin, the mean (i.e., black dot) and standard error (i.e., error bar) of the dependent variable are shown. The x-axis is in log scale, and the median of the number of close friends in each bin was labeled in the x-axis. The red line is the fitted quadratic model. The names of the brain regions are from the HCP-MMP atlas.

After FDR correction (*q* < 0.05), the significant cortical areas associated with the number of close friends were mainly located in the OFC, insula, the ACC, the anterior temporal cortex, and the TPJ (Figure 3a and Table S3). The brain region with the largest effect size for the linear term was the OFC (left medial OFC [area 11l and 13l] and lateral OFC [area 47m and 47s]). The quadratic terms of the number of close friends for all these regions were significant (Figure 3b), and the greatest effect sizes were observed in the temporal pole (left STGa: *β* = -0.001, *t* = -4.20, *p* = 2.8⨉10^−5^, ΔR^2^ = 0.18%, Figure 3e; right TGd: *β* = -0.007, *t* = -4.11, *p* = 4.0⨉10^−5^, ΔR^2^ = 0.18%, Figure 3f). These findings were robust for random choice of the siblings (Figure S6). Similar findings were found for cortical volumes (Table S4 and Figure S7). As the correlation of cortical area and cortical volume with the number of close friends is high (r = 0.78, *p* = 3.3×10^−76^) and cortical area and volume themselves are highly correlated (r = 0.92, *p* = 9.0×10^−151^; Figure S8), we focused on cortical area in the following analyses.

Further, two-lines tests suggested that participants with around 5 close friends (breakpoint = 5.30 ± 0.85) had the largest areas in these cortical regions (Figure S9). To illustrate the patterns of nonlinear relationships, we performed linear regression models in participants with ≤ 5 and > 5 close friends, respectively. Similar regions to those found with quadratic models including the OFC, insula, the ACC, and temporal cortex were significant after FDR correction in the ≤ 5 group (Figure S10a and S10b), and the largest effect size was observed in the OFC (Figure S10c). However, the number of close friends was not related to cortical area in the > 5 group (Figure S10d).

The differences in cortical area related to the number of close friends in the two groups were not correlated (r = -0.02, *p* = 0.78; Figure S10e).

### Relationship to molecular architecture

As the number of close friends was nonlinearly associated with cortical area and the significant regions were only found in participants with no more than 5 close friends, we focused on the brain difference pattern for the number of close friends in the ≤ 5 group. We found that the correlations between the spatial pattern of cortical area related to the number of close friends and densities of neurotransmitters were not significant except for the μ-opioid receptor (Spearman’s rho = 0.44, Bonferroni corrected *p*_perm_ = 0.02; Figure 4a and 4b). Transcriptomic analyses showed that OPRM1 (Spearman’s rho = 0.45, *p*_perm_ = 0.001; Figure 4c) and OPRK1 (Spearman’s rho = 0.46, *p*_perm_ = 0.002; Figure 4d) were highly expressed in regions related to the number of close friends.

**Figure 4.**
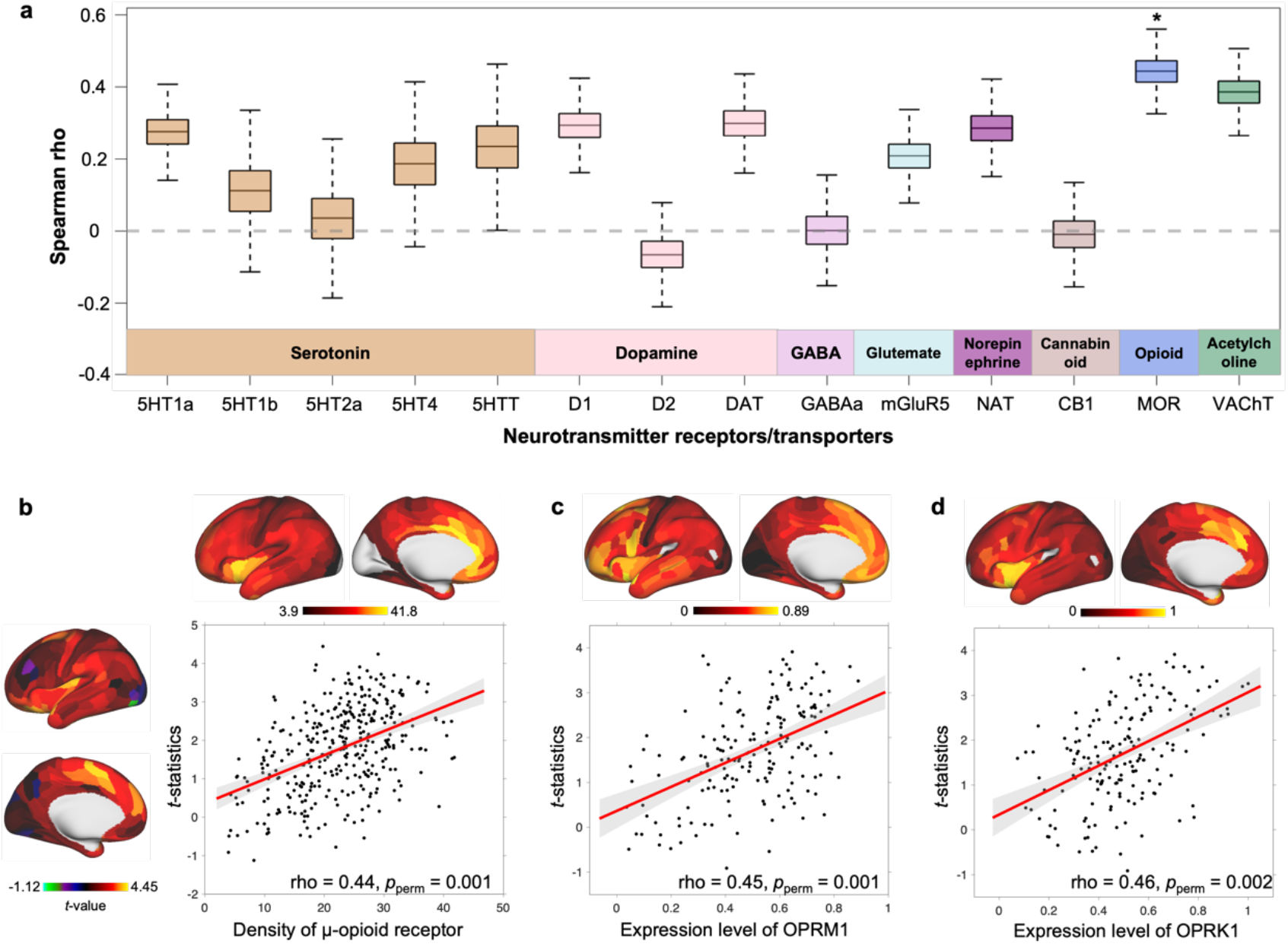
Spatial correlation between cortical area differences related to the number of close friends in children with ≤ 5 close friends and density of neurotransmitters and gene expression level. Bootstrapped Spearman correlations (10,000 times) between *t*-statistics of close friendship quantity and densities of 14 neurotransmitter receptors or transporters (a). In each box, the line indicates the median and the whiskers indicate the 5th and 95th percentiles. *P* values were estimated by 5,000 times permutation. *: Bonferroni corrected *p*_perm_ < 0.05. MOR: μ-opioid receptor. The scatter map of *t*-statistics of close friendship quantity and the density of μ-opioid receptor (b). The scatter map of *t*-statistics of close friendship quantity and the expression level of OPRM1 (c). The scatter map of *t*-statistics of close friendship quantity and the expression level of OPRK1 (d).

### Longitudinal and mediation results

As the nonlinear association between the number of close friends and ADHD symptoms is relatively strong and robust, and for cognitive outcomes, only crystalized intelligence was collected at 2-year follow-up in the ABCD study, we focused on these two measures in longitudinal and mediation analyses. The cross-lagged panel model (CLPM) revealed that participants having closer to 5 close friends had fewer ADHD symptoms 2 years later (*β* = 0.04, *p* < 0.001; Figure 5a). CLPMs in separate groups confirmed that more close friends contributed to fewer ADHD symptoms in ≤ 5 group (*β* = -0.04, *p* = 0.003; Figure S11a), but the effect reversed in the > 5 group (*β* = 0.05, *p* = 0.019; Figure S11b). The relationship between the absolute difference of close friend number to 5 and crystalized intelligence was bidirectional (Figure 5b). Only in the ≤ 5 group was a significant negative correlation found between crystalized intelligence at baseline and the number of close friends at 2-year follow-up (*β* = -0.06, *p* = 0.001; Figure S11c and S11d).

**Figure 5.**
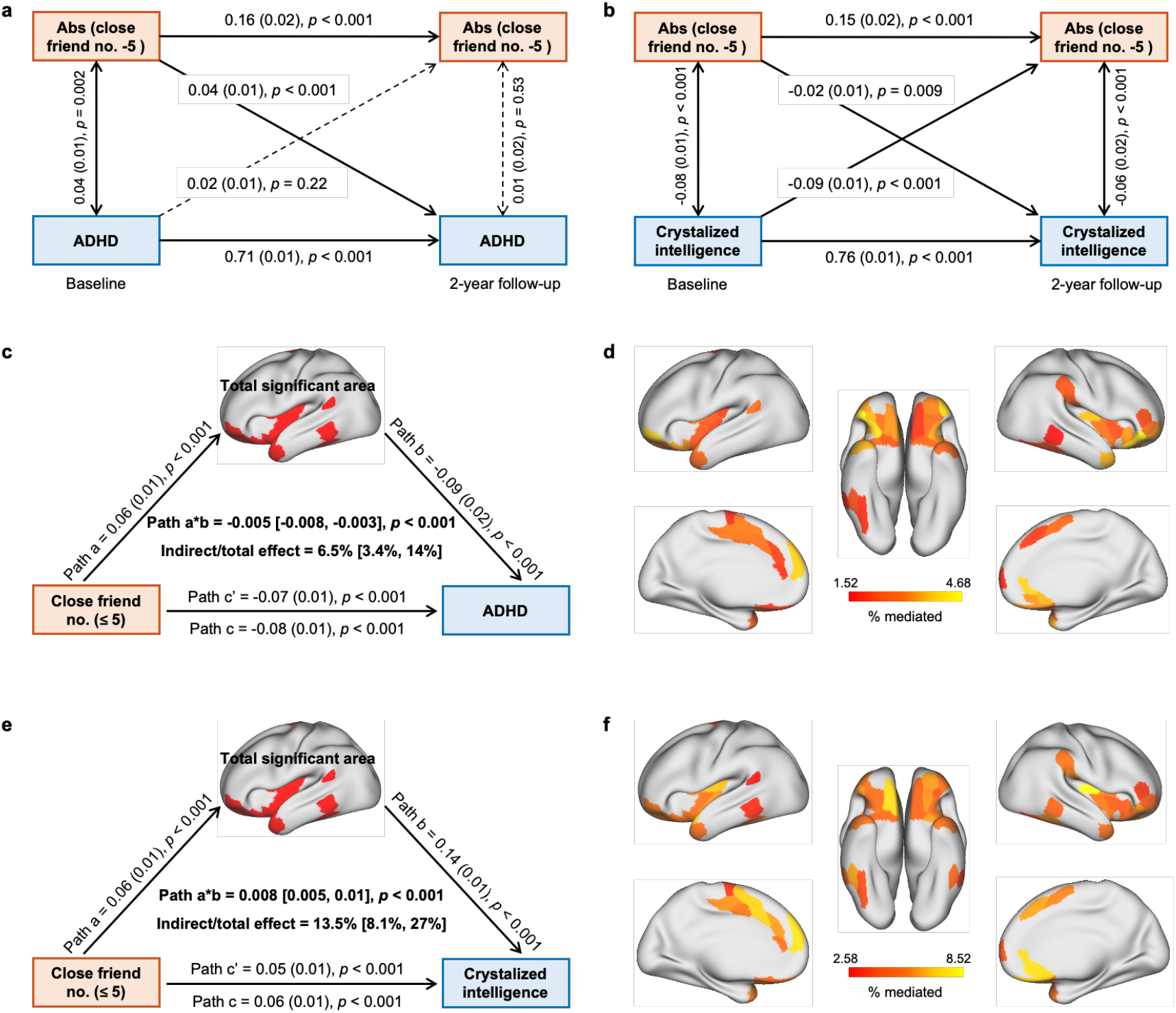
Results of longitudinal and mediation analysis in the ABCD study. CLPM of the absolute value of close friendship quantity to 5 and ADHD symptoms (a). CLPM of the absolute value of close friendship quantity to 5 and crystalized intelligence (b). Mediation analysis of close friendship quantity, the total area of significant regions and ADHD symptoms (c). The effect of individual significant cortical areas that mediated the association between close friendship quantity and ADHD symptoms (d). Mediation analysis of close friendship quantity, the total area of significant regions and crystalized intelligence (e). The effect of individual significant cortical areas that mediated the association between close friendship quantity and crystalized intelligence (f). FDR correction was performed within the significant regions.

Mediation analyses were used to determine whether and the extent to which the association between the number of close friends, ADHD symptoms, and crystalized intelligence could be explained by the identified cortical areas in the ≤ 5 group. The total identified cortical area partly mediated the association between the number of close friends and ADHD symptoms (6.5%, 95%CI [3.4%, 14%]; path a*b: -0.005, 95%CI [-0.008, -0.003]; Figure 5c), and the mediation effects of individual significant regions ranged from 1.52% to 4.68% (Figure 5d). Similarly, the association between the number of close friends and crystalized intelligence was partly mediated by the total identified cortical area (13.5%, 95%CI [8.1%, 27%]; path a*b: 0.008, 95%CI [0.005, 0.01]; Figure 5e), ranging from 2.58% to 8.52% for each significant region (Figure 5f).

### Findings in an independent social network dataset

Utilizing the social network dataset allowed us to validate and extend findings in the ABCD study, as it is an independent and large dataset, the friendship network was generated by nomination, and different measures of mental health and cognition were collected (i.e., well-being and grade point average [GPA]). Three indicators of friendship network size (i.e., outdegree, indegree, and reciprocal degree; Figure S12) were significantly related to well-being (indegree: F = 38.63, *p* = 1.8×10^−17^; outdegree: F = 33.55, *p* = 2.9×10^−15^; reciprocal degree: F = 53.87, *p* = 4.8×10^−24^; Figure S13a) and GPA (indegree: F = 28.08, *p* = 6.7 × 10^−13^; outdegree: F = 46.66, *p* = 6.2 × 10^−21^; reciprocal degree: F = 192.65, *p* = 2.1×10^−83^; Figure S13b). Specifically, for well-being, all linear terms were significant, but only the quadratic term of outdegree was significant after Bonferroni correction (*β* = -2.9⨉10^−4^, *t* = -3.67, *p* = 2.4⨉10^−4^, ΔR^2^ = 0.07%; Figure S13c). For GPA, the quadratic terms of all three indicators were significant, and the greatest effect size was observed in the outdegree (*β* = -0.001, *t* = - 6.02, *p* = 1.8⨉10^−9^, ΔR^2^ = 0.17%; Figure S13c). The two-lines tests revealed that the positive association of outdegree with well-being and GPA diminished once the outward nomination reached 7 or 8 (Figure S13d). The results confirmed that friendship network size especially outdegree was nonlinearly related to mental health and cognitive outcomes.

## Discussion

The present study showed that close friendship quantity was associated with better mental health and higher cognitive functions in late childhood, however, the beneficial effects diminished or reversed when increasing the number of close friends beyond a moderate level. Consistent with the hypothesis, a quadratic association between the number of close friends and the areas of social brain regions was found, including the orbitofrontal cortex (OFC), the anterior cingulate cortex (ACC), insula, anterior temporal cortex and temporo-parietal junction (TPJ), and these regions mediated the nonlinear association with behavior. In addition, the brain differences related to the number of close friends were correlated with measures of the endogenous opioid involvement of the brain regions.

Applying two different analytic approaches in two independent large-scale datasets, we provide compelling evidence that the number of close friends was nonlinearly associated with various mental health and cognitive outcomes. Too large a social network size or too frequent social contacts were not beneficial for well-being and were even detrimental for physical and mental health^10–12,28^. One explanation is that the quantity of relationships an individual can maintain at the same time is limited by a combination of cognitive capacity and time^8^. There is a trade-off between the quantity and quality of friendships, suggesting that an increasing number of close friends may be associated with less intimacy. Meanwhile, as time resources are finite, spending too much time on social activities may lead to insufficient time for study and thereby to lower academic performance. Indeed, adolescents are susceptible to peer influence^29^. The presence of a peer may increase risk-taking behaviors which can be detrimental to mental health^30^, and reduce cognitive performance^31^. Having more close friends may increase the possibility of this kind of influence. In addition, ADHD symptoms or novelty seeking may lead to searching for many superficial friendships. Further, in the ABCD study, we identified an optimal number of 5 close friends, which is consistent with the Dunbar’s number^9^. People devote about 40% of their total social efforts (e.g., time and emotional capital) to just their 5 most important people^32^. In a phone-call dataset of almost 35 million users and 6 billion calls, a layered structure was found with the innermost layer of an average of 4.1 people^33^. Our study extends previous findings that there may be an optimal size of close friendship network in late childhood in terms of its relationship to mental health, cognition, and brain structure. The number of 5 close relationships appears to be consistent with the social network size limited by cognitive and time resources found in anthropological studies, and greatly extends previous research by relating this to brain systems and transmitter systems in the brain.

Although friendships are recognized to be especially significant for psychosocial development in late childhood and adolescence, we know relatively little about how brain development relates to close friendships^34^. Consistent with the behavioral findings, we found that the number of close friends was nonlinearly related to the cortical areas of social brain regions. Children with more close friends had larger areas of social brain regions, but the positive relationship only held for the group with no more than 5 close friends. The areas of social brain regions partly mediated the relationship of the number of close friends with ADHD symptoms and crystalized intelligence. There are two major systems in the brain related to social behavior: an affective system of the OFC, ACC and the anterior insula, and a mentalizing system including the TPJ^35,36^. The dorsal ACC and anterior insula play an important role in social pain (i.e., painful feelings associated with social disconnection)^37^. The OFC receives information about socially relevant stimuli such as face expression and gesture from the cortex in the superior temporal sulcus^17,38^, and is involved in social behavior by representing social stimuli in terms of their reward value^35,39,40^. The OFC can then influence behavior by a number of routes including the ACC^35,41–43^. The OFC volume is associated with social network size, which is partly mediated by mentalizing competence^44^. Previously published meta-analysis studies reported an overlap in brain activation between all mentalizing tasks in the mPFC and posterior TPJ^45^. Stimulation of the TPJ improves self-other representations^46^. Animal studies confirmed a casual effect of social relationships on brain development. Adolescent rodents with deprivation of peer contacts showed brain level changes including reduced synaptic pruning in the prefrontal cortex^47^.

It is known that the endogenous opioid system has an important role in social affiliative processes^22^. In vivo human positron emission tomography (PET) studies found that μ-opioid receptor regulation in brain regions such as the amygdala, anterior insula, and the ACC may preserve and promote emotional well-being in the social environment^48^. Variation in the μ-opioid receptor gene (OPRM1) was associated with individual differences in rejection sensitivity, which was mediated by dorsal ACC activity in social rejection^21^. OPRM1 variation was also related to social hedonic capacity^49^. Pain tolerance, which is associated with activation of the μ-opioid receptor, was correlated with social network size in humans^50^. Social behaviors like social laughter and social touch increased pleasurable sensations and triggered endogenous opioid release to maintain social relationships^51–53^. The opioid system is associated with major psychiatric disorders especially depression^54^.

Several issues should be taken into account when considering our findings. First, as an association study, no causal conclusion should be made in this study. Whether the number of close friends drives the social brain development, or children with larger social brains have more close friends, is unclear. A bidirectional relationship has been reported in the literature^14^. Second, the egocentric network size used in the ABCD study is not able to differentiate reciprocal and unilateral friendships. It is possible that the findings may differ using different indicators^55,56^. For example, in the social network dataset, we showed that outdegree but not indegree and reciprocal degree was quadratically associated with the GPA. Third, the quality of close friendships has not been considered in this study. It has been reported that the relationship between having more friends and fewer depressive symptoms in adolescence was mediated by a sense of belonging^57^. Although current findings on the relative importance of friendship quantity and quality are inconsistent^58,59^, it is necessary to include their interactions in future studies. Finally, although we found children with 5 close friends had the best mental health and cognitive functions, and the largest social brain areas, the optimal size cannot be generalized to populations of other ages and cultures. Extension of the findings in other cultural and age groups would be useful.

In conclusion, this study provides new evidence going beyond previous research that a larger number of close friends up to a moderate level in late childhood is associated with better mental health and higher cognitive functions, and that this can be partly explained by the size of the social brain including the OFC and ACC, and the endogenous opioid system. This study may have implications for targeted friendship interventions in the transition from late childhood to early adolescence.

## Materials and Methods

### Participants and behavioral measures

#### The ABCD study

The Adolescent Brain Cognitive Development (ABCD) Study is tracking the brain development and health of a nationally-representative sample of children aged 9 to 11 years from 21 centers throughout the United States (https://abcdstudy.org). Parents’ full written informed consent and all children’s assent were obtained by each center, and research procedures and ethical guidelines were followed in accordance with the Institutional Review Boards^60^. The current study was conducted on the ABCD Data Release 4.0. At baseline, 8,835 individuals from 7,512 families (6,225 [82.9%] with a child, 1,252 [16.7%] with 2 children, 34 [0.5%] with 3 children, and 1 [0.01%] with 4 children) had complete behavioral and structural MRI data. To avoid the influence of family relatedness, we randomly picked only one child in each family, finally resulting in 7,512 children, of whom 4,290 had two-year follow-up data.

Close friendships are characterized by enjoying spending time together, having fun, and trust. Participants were asked how many close friends that are boys and girls they have, respectively. Mental health problems were rated by the parent using the Child Behavior Checklist (CBCL), a validated and widely-used assessment of childhood behavior ^61^. The CBCL contains 20 empirically based subscales spanning emotional, social and behavioral domains. Raw scores were used in analyses, higher scores indicating more severe problems. Cognitive functions were assessed by the NIH Toolbox®^62^, which has good reliability and validity in children^63^. The toolbox consists of seven different tasks covering episodic memory, executive function, attention, working memory, processing speed, and language abilities, and also provides three composites of crystalized, fluid, and total intelligence^64^. Uncorrected standard scores were used in analyses. All ten cognitive scores were available at baseline, but only crystalized intelligence was collected two years later.

#### Social network dataset

In order to validate and generalize the findings in the ABCD study, we utilized a publicly available dataset of a social network experiment, conducted among students in 56 middle schools in New Jersey, the United States^26^ (https://www.icpsr.umich.edu/web/civicleads/studies/37070). All parents and students provided informed consent for the survey, and the research protocol was approved by the Princeton University Institutional Review Board. Participants were asked to report which other students (up to ten) in their school they chose to spend time with in the last few weeks. We were able to generate a directed friendship network within each school, and therefore identified three kinds of network measures for each individual: outdegree (i.e., outward nomination), indegree (i.e., inward nomination), and reciprocal degree (i.e., reciprocal nomination). Well-being was assessed by three questions: “I feel like I belong at this school”, “I have stayed home from school because of problems with other students”, and “During the past month, I have often been bothered by feeling sad and down”^11^. Cognitive function was measured by the GPA on a 4.0 scale, obtained from school administrative records.

#### Structural MRI data

In the ABCD study, 3D T1- and T2-weighted structural images were collected using 3T scanners at 21 data collecting sites^65^. The detailed preprocessing pipeline has been described elsewhere^66^. In brief, we used FreeSurfer v6.0 to preprocess the minimal preprocessed T1- and T2-weighted images downloaded from the ABCD study, including cortical surface reconstruction, subcortical segmentation, smoothed by a Gaussian kernel (FWHM = 10 mm), and estimation of morphometric measures (i.e., cortical area, thickness, and volume). Then, the cortical surface of each subject was registered to a standard fsaverage space and parcellated into 180 cortical regions per hemisphere as defined in the Human Connectome Project multimodal parcellation (HCP-MMP) atlas^67^. Volumetric reconstructions of subcortical structures were also obtained based on the Aseg atlas^68^.

#### Neurochemical data

Fourteen receptors and transporters across eight different neurotransmitter systems (serotonin: 5HT1a, 5HT1b, 5HT2a, 5HT4, and 5HTT; dopamine: D1, D2, and DAT; GABA: GABAa; glutamate: mGluR5; norepinephrine: NAT; cannabinoid: CB1; opioid: MOR; acetylcholine: VAChT) were investigated. Density estimates were derived from average group maps of healthy volunteers scanned in prior PET and SPECT studies (Table S5). All density maps were downloaded online (https://github.com/juryxy/JuSpace/tree/JuSpace_v1.3/JuSpace_v1.3/PETatlas), which had been registered and normalized into the Montreal Neurological Institute (MNI) space, and linearly rescaled to 0-100^69^. For comparability, the HCP atlas in fsaverage space was converted to individual surface space (“mri_surf2surf”) of the MNI brain template ch2^70^ which was preprocessed by Freesurfer (“recon-all”), and then was projected to volume (“mri_label2vol”). The density maps were parcellated into the 360 cortical regions as the structural MRI data according to the volume-based HCP-MMP atlas. Specifically, for the μ-opioid receptor, occipital cortex served as the reference region^71^, and was therefore excluded in analysis.

#### Transcriptomic data

Gene expression data was from six neurotypical adult brains in the Allen Human Brain Atlas^72^. We focused on the opioid receptor genes (i.e., OPRM1 and OPRK1). The preprocessed transcriptomic data were imported from 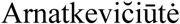 et al^73^ (https://doi.org/10.6084/m9.figshare.6852911), including probe-to-gene re-annotation, intensity-based data filtering, and probe selection using RNA-seq data as a reference. Then, samples were assigned to brain regions according to the volume-based HCP-MMP atlas, and expression values were averaged within each region. Since right hemisphere data were only available for two donors, analyses were conducted on the left hemisphere only, finally resulting in 177 brain regions.

## Statistical analysis

### Nonlinear association analysis

The nonlinear associations of close friendship quantity with mental health, cognition, and brain structure were investigated. The quantity of close friendship was log-transformed [log_10_(x+1)] in analyses as it has a skewed distribution^55^ (Figure S14). Two different analytic approaches were used to robustly evaluate the nonlinear relationships. First, we fitted a quadratic regression model (y = bx^2^ + ax + c) with close friendship quantity as the independent variable. Close friendship quantity was mean-centered to ensure that the linear (a) and quadratic (b) terms were orthogonal. Three statistical parameters were of interest: a total F-value of linear and quadratic terms, reflecting the association between close friendship quantity and the measures of interest^74^; the quadratic term, indicating the presence of a nonlinear association; and the linear term. The effect size of the quadratic term was calculated by the change in the overall proportion of variance (adjusted R^2^) between the quadratic model and the corresponding linear model, and the effect size of the linear term was the ΔR^2^ between the linear model and the model with only covariates. The model fits of quadratic and linear models were compared by analysis of variance. Although quadratic regression is widely used in psychosocial studies to detect the presence of nonlinearity^11,75^, simulation studies showed that this approach for testing a U-shaped effect has a high false positive rate^27^. Therefore, we conducted a two-lines test^27^ once a significant quadratic term was found, which could estimate a data-driven breakpoint. We then split the data accordingly to fit two linear models, respectively. If the segment slopes have opposite signs and both of them are significant, a U-shaped relation exists. Same analytic approaches were used in behavioral and neuroimaging analyses. In the ABCD study, sex, age, parent education level, household income, ethnicity, puberty, BMI and site were used as covariates of no interest for the behavioral analyses. For the neuroimaging analyses, we additionally controlled for handedness, head motion and MRI manufacturer. In the social network dataset, we controlled for sex, age, grade, whether the subject was or was not new to the school, and whether or not most friends went to this school. Bonferroni correction was used in behavioral analyses, and FDR correction was used in neuroimaging analyses.

Several sensitivity analyses were performed. To examine the potential sex influence, we conducted nonlinear association analyses in male and female, respectively. The effect of the sex of close friends was tested by separating close friends into same-sex and opposite-sex ones. To validate the findings from data at baseline, we replicated the same analyses using the cross-sectional data collected at 2 years later. For neuroimaging analyses, if significant nonlinear associations were detected, we also conducted linear regression models in two groups split by the average breakpoint, respectively.

### Spatial correlation with neurotransmitter density and gene expression

Unthresholded *t*-statistic maps of brain structure associated with close friendship quantity in two groups (i.e., split by the average breakpoint) were used to correlate with neurotransmitter density and gene expression level by Spearman’s rank correlation. Bootstrapping was performed to ensure the robustness, and the significance was tested by 5,000 times permutation, in which the correlation was re-computed using null *t*-statistic maps obtained by label shuffling for close friendship quantity^76^.

### Cross-lagged panel analysis

Longitudinal relationships of close friendship quantity with ADHD symptoms (i.e., cbcl_scr_syn_attention) and crystalized intelligence was investigated using a classic two-wave cross-lagged panel model (CLPM) implemented by Mplus 7.0. Firstly, we conducted CLPMs using the absolute value of the difference between close friendship quantity and the breakpoint, and then established CLPMs for participants with the quantity of close friendship ≤ breakpoint and > breakpoint at baseline, respectively. We controlled for several stable (i.e., sex, parent education level, ethnicity, and site) and time-variant variables (i.e., age, household income, and puberty) in these models. The model parameters were estimated by the full information maximum likelihood method^77^, and the model fit was interpreted using common thresholds of good fit^78^.

### Mediation analysis

We used the baseline data in the ABCD study to test the associations between close friendship quantity, ADHD symptoms, crystalized intelligence, and brain structure. The total area of the significant brain regions was used as the mediator. Variables were normalized and then entered into the model. Sex, age, parent education level, household income, ethnicity, pubertal status, BMI, handedness, head motion, MRI manufacturer and site were used as covariates of no interest. In addition to the total area, the mediation effect of individual significant regions was also evaluated, the *p*-values of the mediation effect corrected by FDR correction. Total, direct and indirect associations were estimated by bootstrapping 10,000 times, and the 95% bias-corrected and accelerated confidence interval (CI) was reported. Analyses were performed using the R mediation package.

## Data Availability

The ABCD data used in this study is Data Release 4.0 (http://dx.doi.org/10.15154/1523041). The data are available by request from the NIMH Data Archive (https://data-archive.nimh.nih.gov/abcd). The social network dataset is conducted among students in 56 middle schools in New Jersey, the United States. This dataset is publicly available (https://www.icpsr.umich.edu/web/civicleads/studies/37070).

## Acknowledgements

We thank the children and families whose ongoing participation made this study possible. Data used in the preparation of this article were obtained from the Adolescent Brain Cognitive Development^SM^ (ABCD) Study (https://abcdstudy.org), held in the NIMH Data Archive (NDA). This is a multisite, longitudinal study designed to recruit more than 10,000 children age 9-10 and follow them over 10 years into early adulthood. The ABCD Study® is supported by the National Institutes of Health and additional federal partners under award numbers U01DA041048, U01DA050989, U01DA051016, U01DA041022, U01DA051018, U01DA051037, U01DA050987, U01DA041174, U01DA041106, U01DA041117, U01DA041028, U01DA041134, U01DA050988, U01DA051039, U01DA041156, U01DA041025, U01DA041120, U01DA051038, U01DA041148, U01DA041093, U01DA041089, U24DA041123, U24DA041147. A full list of supporters is available at https://abcdstudy.org/federal-partners.html. A listing of participating sites and a complete listing of the study investigators can be found at https://abcdstudy.org/consortium_members/. ABCD consortium investigators designed and implemented the study and/or provided data but did not necessarily participate in the analysis or writing of this report. This manuscript reflects the views of the authors and may not reflect the opinions or views of the NIH or ABCD consortium investigators. C.S. is supported by grants from the National Natural Sciences Foundation of China (No. 82101617). W.C. is supported by grants from the National Natural Sciences Foundation of China (No. 82071997), the Shanghai Rising-Star Program (No. 21QA1408700). J.F. is supported by National Key R&D Program of China (No. 2018YFC1312904 and No. 2019YFA0709502), Shanghai Municipal Science and Technology Major Project (No. 2018SHZDZX01), ZJ Lab, Shanghai Center for Brain Science and Brain-Inspired Technology, and the 111 Project (No. B18015).

## References

1. Blakemore, S.-J. & Mills, K. L. Is Adolescence a Sensitive Period for Sociocultural Processing? Annu. Rev. Psychol. 65, 187–207 (2014).

2. Mills, K. L., Lalonde, F., Clasen, L. S., Giedd, J. N. & Blakemore, S.-J. Developmental changes in the structure of the social brain in late childhood and adolescence. Soc Cogn Affect Neurosci 9, 123–31 (2014).

3. Kessler, R. C. et al. Lifetime Prevalence and Age-of-Onset Distributions of DSM-IV Disorders in the National Comorbidity Survey Replication. Arch. Gen. Psychiatry 62, 593–602 (2005).

4. Paus, T., Keshavan, M. & Giedd, J. N. Why do many psychiatric disorders emerge during adolescence? Nat. Rev. Neurosci. 9, 947–957 (2008).

5. Narr, R. K., Allen, J. P., Tan, J. S. & Loeb, E. L. Close Friendship Strength and Broader Peer Group Desirability as Differential Predictors of Adult Mental Health. Child Dev. 90, 298–313 (2019).

6. Marion, D., Laursen, B., Zettergren, P. & Bergman, L. R. Predicting Life Satisfaction During Middle Adulthood from Peer Relationships During Mid-Adolescence. J. Youth Adolesc. 42, 1299–1307 (2013).

7. Wentzel, K. R., Jablansky, S. & Scalise, N. R. Do Friendships Afford Academic Benefits? A Meta-analytic Study. Educ. Psychol. Rev. 30, 1241–1267 (2018).

8. Dunbar, R. I. M. The Anatomy of Friendship. Trends Cogn. Sci. 22, 32–51 (2018).

9. Zhou, W.-X., Sornette, D., Hill, R. A. & Dunbar, R. I. M. Discrete hierarchical organization of social group sizes. Proc. R. Soc. B Biol. Sci. 272, 439–444 (2005).

10. Falci, C. & McNeely, C. Too Many Friends: Social Integration, Network Cohesion and Adolescent Depressive Symptoms. Soc. Forces 87, 2031–2061 (2009).

11. Ren, D., Stavrova, O. & Loh, W. W. Nonlinear effect of social interaction quantity on psychological well-being: Diminishing returns or inverted U? J. Pers. Soc. Psychol. 122, 1056–1074 (2022).

12. Kushlev, K., Heintzelman, S. J., Oishi, S. & Diener, E. The declining marginal utility of social time for subjective well-being. J. Res. Personal. 74, 124–140 (2018).

13. Brown, M. I., Wai, J. & Chabris, C. F. Can You Ever Be Too Smart for Your Own Good? Comparing Linear and Nonlinear Effects of Cognitive Ability on Life Outcomes. Perspect. Psychol. Sci. 23.

14. Dunbar, R. I. M. & Shultz, S. Evolution in the Social Brain. Science 317, 1344–1347 (2007).

15. Sallet, J. et al. Social Network Size Affects Neural Circuits in Macaques. Science 334, 697–700 (2011).

16. Testard, C. et al. Social connections predict brain structure in a multidimensional free-ranging primate society. Sci. Adv. 8, eabl5794 (2022).

17. Pitcher, D. & Ungerleider, L. G. Evidence for a third visual pathway specialized for social perception. Trends Cogn. Sci. 25, 100–110 (2021).

18. Blakemore, S.-J. The social brain in adolescence. Nat. Rev. Neurosci. 9, 267–277 (2008).

19. Burnett, S., Sebastian, C., Cohen Kadosh, K. & Blakemore, S.-J. The social brain in adolescence: Evidence from functional magnetic resonance imaging and behavioural studies. Neurosci. Biobehav. Rev. 35, 1654–1664 (2011).

20. Baumgärtner, U. et al. High opiate receptor binding potential in the human lateral pain system. NeuroImage 30, 692–699 (2006).

21. Way, B. M., Taylor, S. E. & Eisenberger, N. I. Variation in the μ-opioid receptor gene (OPRM1) is associated with dispositional and neural sensitivity to social rejection. Proc. Natl. Acad. Sci. 106, 15079–15084 (2009).

22. Machin, A. J. & Dunbar, R. I. M. The brain opioid theory of social attachment: a review of the evidence. Behaviour 148, 985–1025 (2011).

23. Porcelli, S. et al. Social brain, social dysfunction and social withdrawal. Neurosci. Biobehav. Rev. 97, 10–33 (2019).

24. Lamblin, M., Murawski, C., Whittle, S. & Fornito, A. Social connectedness, mental health and the adolescent brain. Neurosci. Biobehav. Rev. 80, 57–68 (2017).

25. Karcher, N. R. & Barch, D. M. The ABCD study: understanding the development of risk for mental and physical health outcomes. Neuropsychopharmacology 46, 131–142 (2021).

26. Paluck, E. L., Shepherd, H. & Aronow, P. M. Changing climates of conflict: A social network experiment in 56 schools. Proc. Natl. Acad. Sci. 113, 566–571 (2016).

27. Simonsohn, U. Two Lines: A Valid Alternative to the Invalid Testing of U-Shaped Relationships With Quadratic Regressions. Adv. Methods Pract. Psychol. Sci. 1, 538–555 (2018).

28. Stavrova, O. & Ren, D. Is More Always Better? Examining the Nonlinear Association of Social Contact Frequency With Physical Health and Longevity. Soc. Psychol. Personal. Sci. 12, 1058–1070 (2021).

29. Berndt, T. J. Developmental changes in conformity to peers and parents. Dev. Psychol. 15, 608–616 (1979).

30. Chein, J., Albert, D., O’Brien, L., Uckert, K. & Steinberg, L. Peers increase adolescent risk taking by enhancing activity in the brain’s reward circuitry. Dev. Sci. 14, F1–F10 (2011).

31. Wolf, L. K., Bazargani, N., Kilford, E. J., Dumontheil, I. & Blakemore, S.-J. The audience effect in adolescence depends on who’s looking over your shoulder. J. Adolesc. 43, 5–14 (2015).

32. Bzdok, D. & Dunbar, R. I. M. The Neurobiology of Social Distance. Trends Cogn. Sci. 24, 717–733 (2020).

33. Mac Carron, P., Kaski, K. & Dunbar, R. Calling Dunbar’s numbers. Soc. Netw. 47, 151–155 (2016).

34. Pfeifer, J. H. & Allen, N. B. Puberty Initiates Cascading Relationships Between Neurodevelopmental, Social, and Internalizing Processes Across Adolescence. Biol. Psychiatry 89, 99–108 (2021).

35. Rolls, E. T. The Orbitofrontal Cortex. (Oxford University Press, 2019).

36. Schmälzle, R. et al. Brain connectivity dynamics during social interaction reflect social network structure. Proc. Natl. Acad. Sci. 114, 5153–5158 (2017).

37. Eisenberger, N. I. The pain of social disconnection: examining the shared neural underpinnings of physical and social pain. Nat. Rev. Neurosci. 13, 421–434 (2012).

38. Hasselmo, M. E., Rolls, E. T. & Baylis, G. C. The role of expression and identity in the face-selective responses of neurons in the temporal visual cortex of the monkey. Behav. Brain Res. 32, 203–218 (1989).

39. Rolls, E. T. The orbitofrontal cortex and emotion in health and disease, including depression. Neuropsychologia 128, 14–43 (2019).

40. Rolls, E. T., Critchley, H. D., Browning, A. S. & Inoue, K. Face-selective and auditory neurons in the primate orbitofrontal cortex. Exp. Brain Res. 170, 74–87 (2006).

41. Rolls, E. T., Cheng, W. & Feng, J. The orbitofrontal cortex: reward, emotion and depression. Brain Commun. 2, fcaa196 (2020).

42. Rolls, E. T., Deco, G., Huang, C.-C. & Feng, J. The human orbitofrontal cortex, vmPFC, and anterior cingulate cortex effective connectome: emotion, memory, and action. Cereb. Cortex bhac070 (2022) doi:10.1093/cercor/bhac070.

43. Eisenberger, N. I., Lieberman, M. D. & Williams, K. D. Does Rejection Hurt? An fMRI Study of Social Exclusion. Science 302, 290–292 (2003).

44. Powell, J., Lewis, P. A., Roberts, N., García-Fiñana, M. & Dunbar, R. I. M. Orbital prefrontal cortex volume predicts social network size: an imaging study of individual differences in humans. Proc. R. Soc. B Biol. Sci. 279, 2157–2162 (2012).

45. Schurz, M., Radua, J., Aichhorn, M., Richlan, F. & Perner, J. Fractionating theory of mind: A meta-analysis of functional brain imaging studies. Neurosci. Biobehav. Rev. 42, 9–34 (2014).

46. Santiesteban, I., Banissy, M. J., Catmur, C. & Bird, G. Enhancing Social Ability by Stimulating Right Temporoparietal Junction. Curr. Biol. 22, 2274–2277 (2012).

47. Orben, A., Tomova, L. & Blakemore, S.-J. The effects of social deprivation on adolescent development and mental health. Lancet Child Adolesc. Health 4, 634–640 (2020).

48. Hsu, D. T. et al. Response of the μ-opioid system to social rejection and acceptance. Mol. Psychiatry 18, 1211–1217 (2013).

49. Troisi, A. et al. Social hedonic capacity is associated with the A118G polymorphism of the mu-opioid receptor gene (OPRM1) in adult healthy volunteers and psychiatric patients. Soc. Neurosci. 6, 88–97 (2011).

50. Johnson, K. V.-A. & Dunbar, R. I. M. Pain tolerance predicts human social network size. Sci. Rep. 6, 25267 (2016).

51. Dunbar, R. I. M. The social role of touch in humans and primates: Behavioural function and neurobiological mechanisms. Neurosci. Biobehav. Rev. 34, 260–268 (2010).

52. Manninen, S. et al. Social Laughter Triggers Endogenous Opioid Release in Humans. J. Neurosci. 37, 6125–6131 (2017).

53. Nummenmaa, L. et al. Social touch modulates endogenous μ-opioid system activity in humans. NeuroImage 138, 242–247 (2016).

54. Peciña, M. et al. Endogenous opioid system dysregulation in depression: implications for new therapeutic approaches. Mol. Psychiatry 24, 576–587 (2019).

55. Hobbs, W. R., Burke, M., Christakis, N. A. & Fowler, J. H. Online social integration is associated with reduced mortality risk. Proc. Natl. Acad. Sci. 113, 12980–12984 (2016).

56. Kwak, S., Joo, W., Youm, Y. & Chey, J. Social brain volume is associated with in-degree social network size among older adults. Proc. R. Soc. B Biol. Sci. 285, 20172708 (2018).

57. Ueno, K. The effects of friendship networks on adolescent depressive symptoms. Soc. Sci. Res. 34, 484–510 (2005).

58. Bruine de Bruin, W., Parker, A. M. & Strough, J. Age differences in reported social networks and well-being. Psychol. Aging 35, 159–168 (2020).

59. Platt, J., Keyes, K. M. & Koenen, K. C. Size of the social network versus quality of social support: which is more protective against PTSD? Soc. Psychiatry Psychiatr. Epidemiol. 49, 1279–1286 (2014).

60. Auchter, A. M. et al. A description of the ABCD organizational structure and communication framework. Dev. Cogn. Neurosci. 32, 8–15 (2018).

61. Achenbach, T. & Rescorla, L. Manual for the ASEBA school-age forms & profiles: an integrated system of multi-informant assessment. (Burlington, VT: ASEBA, 2001).

62. Luciana, M. et al. Adolescent neurocognitive development and impacts of substance use: Overview of the adolescent brain cognitive development (ABCD) baseline neurocognition battery. Dev. Cogn. Neurosci. 32, 67–79 (2018).

63. Akshoomoff, N. et al. NIH Toolbox Cognitive Function Battery (CFB): Composite Scores of Crystallized, Fluid, and Overall Cognition. Monogr. Soc. Res. Child Dev. 78, 119–132 (2013).

64. Weintraub, S. et al. Cognition assessment using the NIH Toolbox. Neurology 80, S54–S64 (2013).

65. Casey, B. J. et al. The Adolescent Brain Cognitive Development (ABCD) study: Imaging acquisition across 21 sites. Dev. Cogn. Neurosci. 32, 43–54 (2018).

66. Gong, W., Rolls, E. T., Du, J., Feng, J. & Cheng, W. Brain structure is linked to the association between family environment and behavioral problems in children in the ABCD study. Nat. Commun. 12, 3769 (2021).

67. Glasser, M. F. et al. A multi-modal parcellation of human cerebral cortex. Nature 536, 171–178 (2016).

68. Fischl, B. et al. Whole Brain Segmentation: Automated Labeling of Neuroanatomical Structures in the Human Brain. Neuron 33, 341–355 (2002).

69. Dukart, J. et al. JuSpace. Hum. Brain Mapp. 42, 555–566 (2021).

70. Rorden, C. & Brett, M. Stereotaxic Display of Brain Lesions. Behav. Neurol. 12, 191–200 (2000).

71. Kantonen, T. et al. Interindividual variability and lateralization of μ-opioid receptors in the human brain. NeuroImage 217, 116922 (2020).

72. Hawrylycz, M. J. et al. An anatomically comprehensive atlas of the adult human brain transcriptome. Nature 489, 391–399 (2012).

73. Arnatkeviciūte, A., Fulcher, B. D. & Fornito, A. A practical guide to linking brain-wide gene expression and neuroimaging data. NeuroImage 189, 353–367 (2019).

74. Li, Y. et al. The brain structure and genetic mechanisms underlying the nonlinear association between sleep duration, cognition and mental health. Nat. Aging 2, 425–437 (2022).

75. Nook, E. C., Sasse, S. F., Lambert, H. K., McLaughlin, K. A. & Somerville, L. H. The Nonlinear Development of Emotion Differentiation: Granular Emotional Experience Is Low in Adolescence. Psychol. Sci. 29, 1346–1357 (2018).

76. Chen, J. et al. Intrinsic Connectivity Patterns of Task-Defined Brain Networks Allow Individual Prediction of Cognitive Symptom Dimension of Schizophrenia and Are Linked to Molecular Architecture. Biol. Psychiatry 89, 308–319 (2021).

77. Muthén, B., Kaplan, D. & Hollis, M. On structural equation modeling with data that are not missing completely at random. Psychometrika 52, 431–462 (1987).

78. Hu, L. & Bentler, P. M. Cutoff criteria for fit indexes in covariance structure analysis: Conventional criteria versus new alternatives. Struct. Equ. Model. Multidiscip. J. 6, 1–55 (1999).

